# Plasma tissue plasminogen activator and plasminogen activator inhibitor-1 in hospitalized COVID-19 patients

**DOI:** 10.1101/2020.08.29.20184358

**Authors:** Yu Zuo, Mark Warnock, Alyssa Harbaugh, Srilakshmi Yalavarthi, Kelsey Gockman, Melanie Zuo, Jacqueline A. Madison, Jason S. Knight, Yogendra Kanthi, Daniel A. Lawrence

**Author notes:** **Correspondence:** Daniel A. Lawrence. **Disclosures:** Daniel A. Lawrence is a member of the board and holds equity in MDI Therapeutics which is developing therapeutic inhibitors of PAI-1.

## Abstract

Patients with coronavirus disease-19 (**COVID-19**) are at high risk for thrombotic arterial and venous occlusions. However, bleeding complications have also been observed in some patients. Understanding the balance between coagulation and fibrinolysis will help inform optimal approaches to thrombosis prophylaxis and potential utility of fibrinolytic-targeted therapies. 118 hospitalized COVID-19 patients and 30 healthy controls were included in the study. We measured plasma antigen levels of tissue-type plasminogen activator **(tPA**) and plasminogen activator inhibitor-1 (**PAI-1**) and performed spontaneous clot-lysis assays. We found markedly elevated tPA and PAI-1 levels in patients hospitalized with COVID-19. Both factors demonstrated strong correlations with neutrophil counts and markers of neutrophil activation. High levels of tPA and PAI-1 were associated with worse respiratory status. High levels of tPA, in particular, were strongly correlated with mortality and a significant enhancement in spontaneous *ex vivo* clot-lysis. While both tPA and PAI-1 are elevated among COVID-19 patients, extremely high levels of tPA enhance spontaneous fibrinolysis and are significantly associated with mortality in some patients. These data indicate that fibrinolytic homeostasis in COVID-19 is complex with a subset of patients expressing a balance of factors that may favor fibrinolysis. Further study of tPA as a biomarker is warranted.

## INTRODUCTION

The close relationship between COVID-19 and thrombosis is of significant clinical importance. There are increasing reports of venous thromboembolism in COVID-19 patients^1,2^, and arterial thrombosis including strokes and myocardial infarctions have been described^2,3^. Histopathology of lung specimens from patients with severe disease demonstrate fibrin-based occlusion of small vessels^4-6^.

COVID-19 is characterized in most patients by minimum prolongation of activated partial thromboplastin time (**aPTT**) and/or prothrombin time (**PT**), and mild, if any, thrombocytopenia^7,8^ suggesting that it is distinct from traditional descriptions of sepsis-induced coagulopathy^9,10^. There are several (possibly synergistic) mechanisms by which SARS-CoV-2 infection may result in macrovascular and microvascular occlusions including cytokine-mediated activation of leukocytes, endothelium, and platelets; hypoxic vasoconstriction; direct activation of endothelial cells by viral transduction^11^; and potentiation of thrombosis by neutrophil extracellular traps (**NETs**)^12-15^. At the same time, bleeding has been described in some patients with COVID-19. For example, a recent multicenter observation of 400 patients hospitalized with COVID-19 demonstrated an overall bleeding rate of 4.8% and a severe bleeding event (World Health Organization grade 3 or 4) rate of 2.3%^16^.

Fibrinolysis is a tightly controlled process whereby a fibrin-rich thrombus is degraded and remodeled by the protease plasmin^17^. This process is regulated by plasminogen activators and inhibitors with the conversion of plasminogen to plasmin being the end result that supports fibrinolysis^17^. The interplay of plasminogen activators—both tissue-type (**tPA**) and urokinase-type (**uPA**)—and their principal inhibitor, plasminogen activator inhibitor-1 (**PAI-1**), plays a key role in regulating fibrinolytic activity^17^. Impaired fibrinolysis has been suggested among COVID-19 patients, which could further heighten thrombotic risk. This has been evidenced by markedly reduced clot lysis at 30 minutes via thromboelastography (**TEG**) in critically-ill patients with COVID-19^18^. *Ex vivo* evaluation of COVID-19 plasma also noted a prolonged clot lysis time, which was more pronounced among critically-ill COVID-19 patients^19^. Furthermore, a case series demonstrated that 11 of 21 COVID-19 patients who underwent rotational thromboelastometry in an intensive care unit met the criteria for fibrinolytic shutdown; 9 of those 11 patients developed thrombosis during their hospitalization^20^. Elevated PAI-1 levels observed in COVID-19 patients has further suggested impaired fibrinolytic ability^21^. The cause of this fibrinolytic shutdown has yet to be elucidated. Here, we aimed to evaluate the potential roles of tPA and PAI-1 in regulating fibrinolytic homeostasis among COVID-19 patients. Given that both bleeding and clotting have been described in COVID-19, we hypothesized that plasma of some patients would demonstrate fibrinolytic shutdown while plasma of others might present a hyper-fibrinolytic state.

## METHODS

### Human samples

Plasma from 118 patients hospitalized with COVID-19 were used in this study. Blood was collected into EDTA by a trained phlebotomist. After completion of hematological testing ordered by the clinician, the remaining plasma was stored at 4°C for up to 48 hours before it was released to the research laboratory. Samples were immediately divided into aliquots and stored at −80°C until testing. All 118 patients had a confirmed COVID-19 diagnosis based on FDA-approved RNA testing. This study complied with all relevant ethical regulations and was approved by the University of Michigan IRB (HUM00179409). Healthy volunteers were recruited through a posted flyer; exclusion criteria for controls included history of a systemic autoimmune disease, active infection, and pregnancy. The 30 controls included 20 females and 10 males, mean age of 41.7 ± 14.4. All COVID-19 plasma samples were treated with solvent/detergent (0.3% v/v tri-(n-butyl) phosphate and 1% Triton X-100) to inactivate the virus^22^. Control plasma samples were similarly treated with the same solvent/detergent.

### Measurement of PAI-1 and tPA antigen

Total PAI-1 and tPA protein was measured as described ^23^. Briefly, 25 μg of either rabbit anti-human PAI-1 (Molecular Innovations) or mouse anti-human tPA clone 2A153 (Molecular Innovations) was coupled to color-coded superparamagnetic beads. 25 μL of standard or diluted sample and 25 μL coupled beads (4000) were incubated for 2 hours in the dark. 25 μL of 2 μg/mL biotinylated rabbit anti-hPAI-1 or biotinylated rabbit anti-htPA antibody (Molecular Innovations) was added to the plate, followed by incubating with phycoerythrin-conjugated streptavidin. The plate was read with a Luminex® 100 System; the setting was 100 μL sample size and 100 events per well. Levels of PAI-1 and tPA were presented as mean ± standard deviations in the text. Active PAI-1 was detected by the same method but using the human uPA protease coupled to the beads as capture^23^.

### Spontaneous lysis assay

To determine the rate of spontaneous lysis, 40 µL of diluted plasma (1:1 in TBS) was added to a microtiter plate and pre-read at 405 nm. Then, 40 µL of 25 nM alpha human thrombin (Haemtech) and 15 mM CaCl_2_ was added and incubated at 37°C for 30 minutes, and the absorbance was read at 405 nm. Twenty µL of TBS was then added to the plate to prevent clot drying during the extended incubation and the plate was read again after 30 minutes and then at 60-minute intervals up to 8 hours.

### Quantification of S100A8/A9 (calprotectin)

Calprotectin levels were measured with the Human S100A8/S100A9 Heterodimer DuoSet ELISA (DY8226-05, R&D Systems) according to the manufacturer’s instructions.

### Statistical analysis

When two groups were present, normally-distributed data were analyzed by two-sided t test and skewed data were analyzed by Mann-Whitney test or Wilcoxon test. For three or more groups, analysis was by one-way ANOVA or Kruskal-Wallis test with correction for multiple comparisons. Normality was assessed by Shapiro-Wilk test. Correlations were tested by Spearman’s method. Data analysis was with GraphPad Prism software version 8. Statistical significance was defined as p<0.05.

## RESULTS

### Tissue-type plasminogen activator and plasminogen activator inhibitor-1 in COVID-19

Utilizing established Luminex platforms, we measured total PAI-1 and tPA levels (detecting both free and complexed PAI-1 and tPA, respectively) in the plasma of 118 patients hospitalized with COVID-19. We similarly assessed 30 healthy controls whose samples had been banked prior to December 2019. Of the 118 COVID-19 patients, the mean age was 61 with a standard deviation of 17 (range 25-95); 54 were female (46%) (**Table 1**). In our cohort, 42% of patients were supported by mechanical ventilation, 8% were receiving high-flow oxygen, 27% were supported by standard nasal cannula, and 24% were breathing ambient air at the time of sample collection. Markedly elevated levels of both PAI-1 and tPA were detected in patients with COVID-19 as compared with healthy controls (mean ± standard deviation 75 ± 46 vs. 40 ± 42 ng/mL, p<0.0001; and 78 ± 68 vs 2.4 ± 2.6 ng/mL, p<0.0001, respectively **Figure 1A-B**). There was a significant correlation between levels of PAI-1 and tPA among COVID-19 patients (r=0.52, p<0.0001) (**Figure 1C**). In summary, both PAI-1 and tPA are markedly elevated in the plasma of patients hospitalized with COVID-19.

**Table 1:**
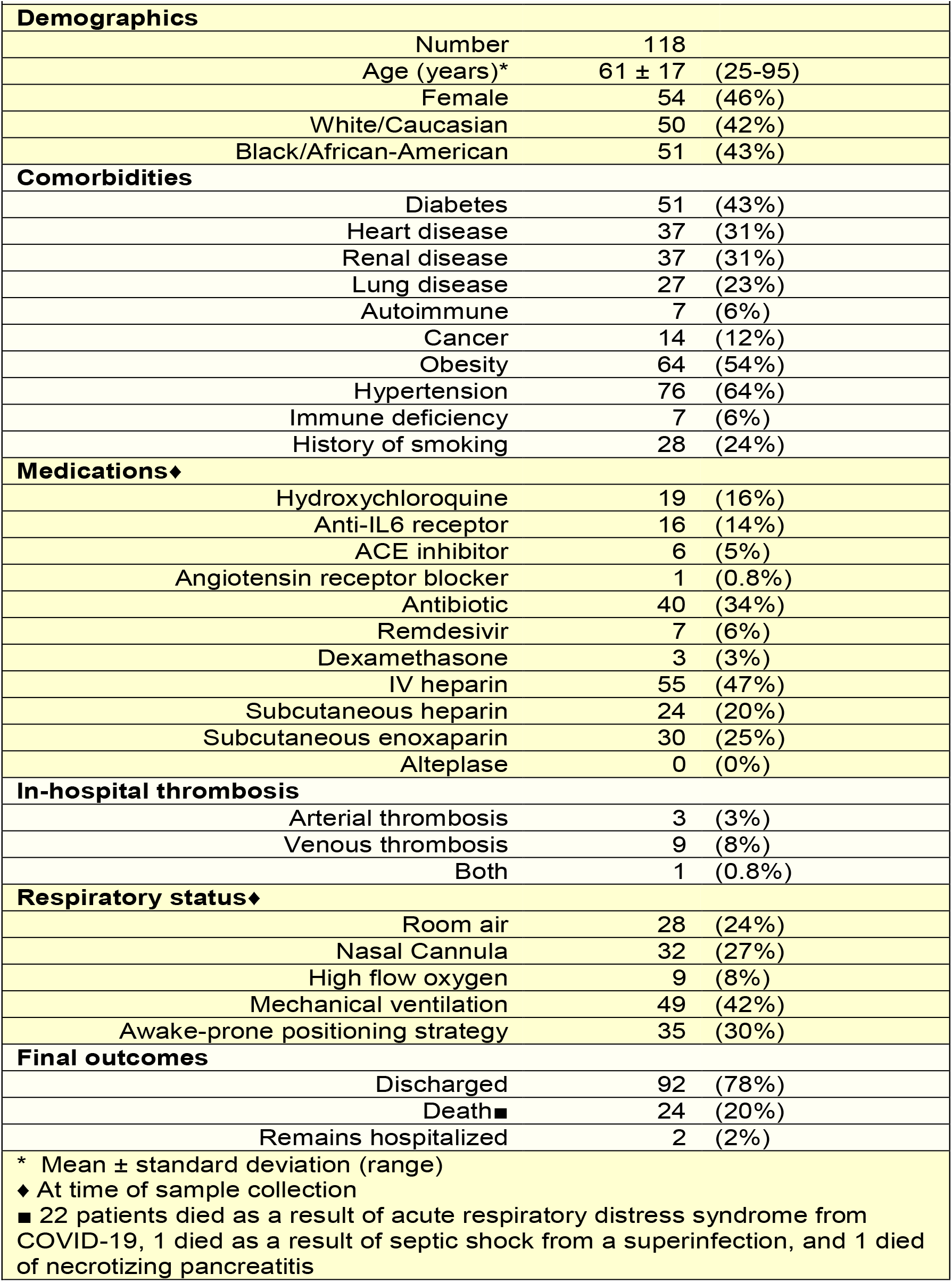
Demographic and clinical characteristics of COVID-19 patients

**Figure 1.**
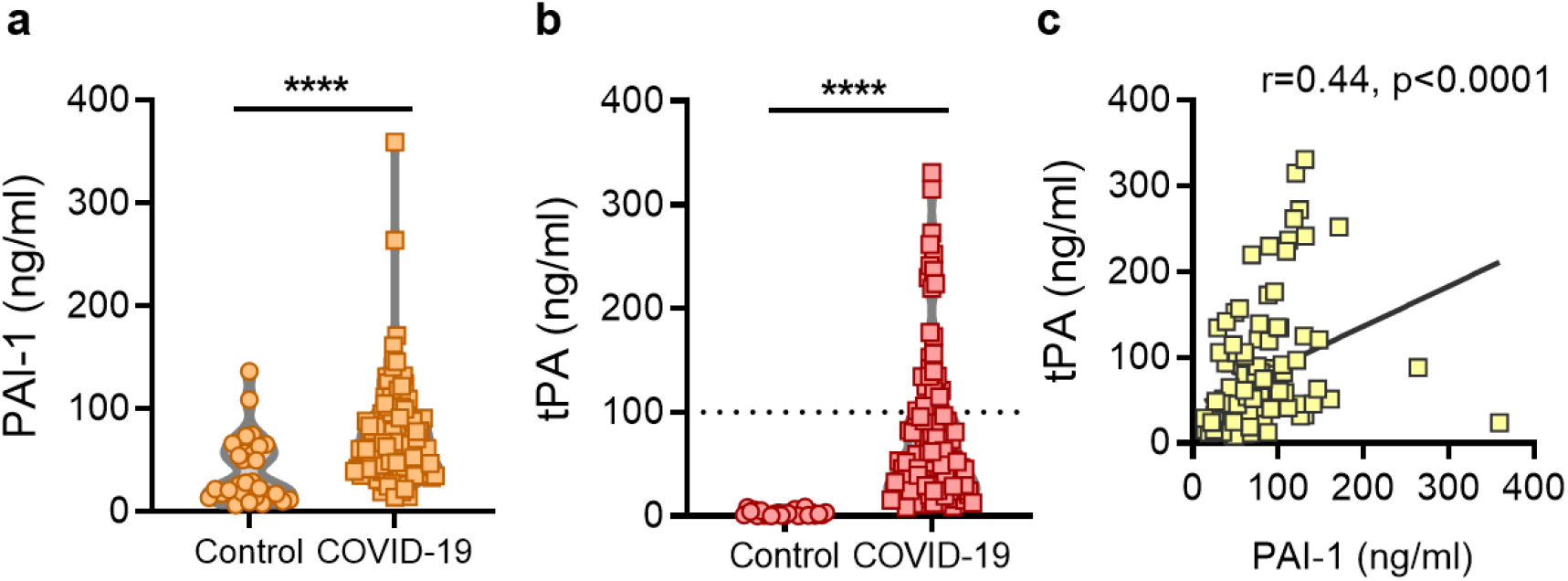
High levels of tPA and PAI-1 among patients with COVID-19. **A-B**, PAI-1 and tPA were measured in individuals with COVID-19, or healthy controls. Levels of PAI-1 and tPA were compared by Mann-Whitney test as samples were not normally distributed when assessed by Shapiro-Wilk test; ****p<0.0001 as compared with the control group. Dotted line indicates high tPA cut-off. **C**, The relationship between tPA and PAI-1 was assessed by Spearman’s correlation test.

### Plasma level of tPA and PAI-1 and their association with clinical biomarkers

We assessed potential correlations with D-dimer and platelet count. We limited the analysis of clinical laboratory measurements to those performed on the same day as plasma used for the PAI-1 and tPA assays. No significant correlation was found between D-dimer and either PAI-1 (r=0.23, p=0.11) or tPA (r=-0.01, p=0.94) (**Figure 2A-B**). We did observe a strong correlation between PAI-1 and platelet count (r=0.33, p=0.0003) (**Figure 2C**); the same was not true for tPA (r=0.06, p=0.5) (**Figure 2D**). Given that activated neutrophils and their products can exert anti-fibrinolytic effects^24^, we next asked how absolute neutrophil count and calprotectin (a marker of neutrophil activation) compared to tPA and PAI-1. Both PAI-1 and tPA demonstrated strong positive correlations with same-day absolute neutrophil count (r=0.32, p=0.03 and r=0.23, p=0.03) (**Figure 2E-F**), as well as levels of calprotectin (r=0.42, p<0.0001 and r=0.23, p=0.01) (**Figure 2G-H**). In summary, levels of PAI-1 and tPA demonstrated strong correlations with neutrophil numbers and activation.

**Figure 2:**
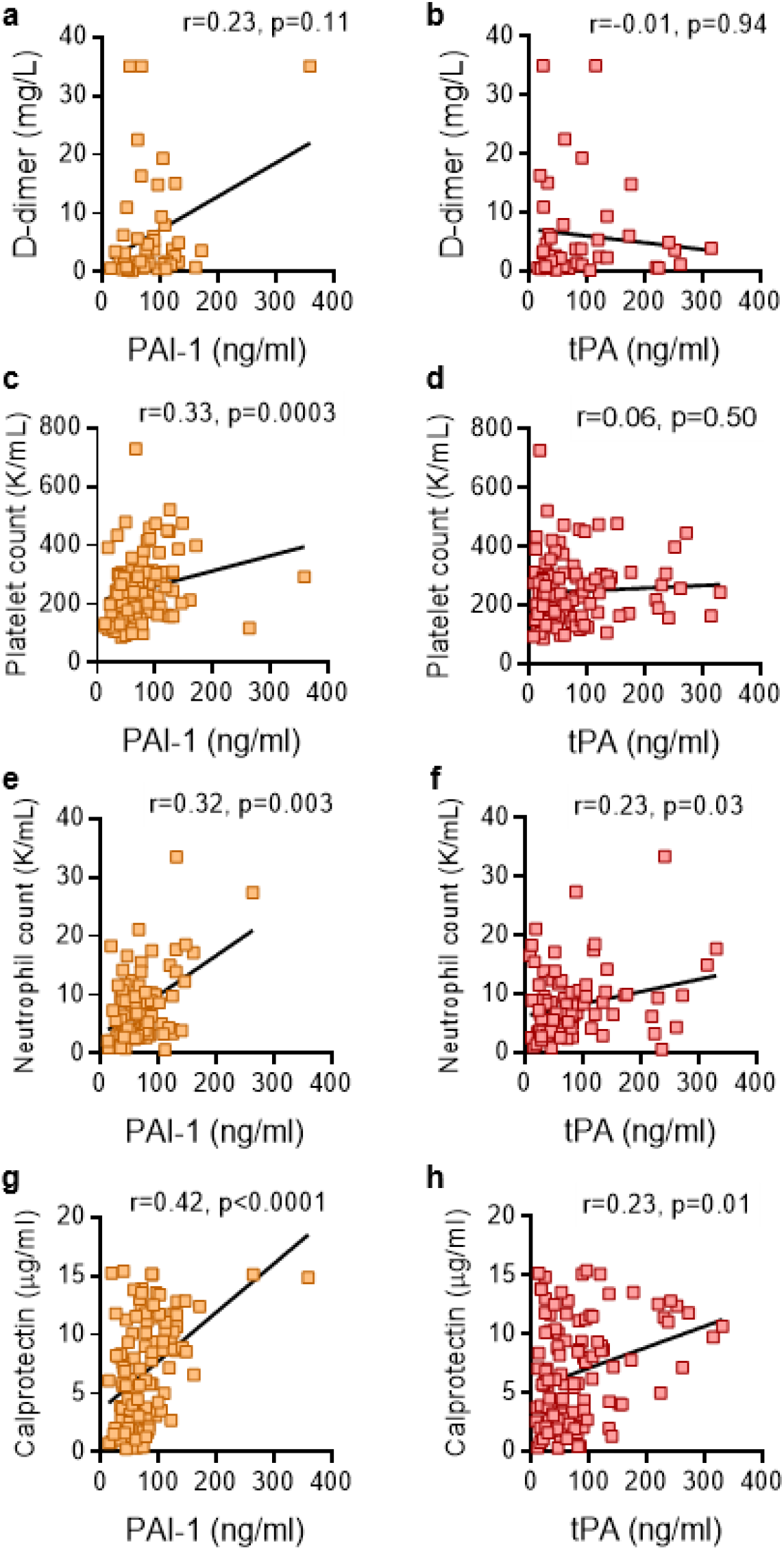
Association between PAI-1 and tPA and clinical biomarkers in plasma. Levels of PAI-1 and tPA were compared to D-dimer (**A, B**), platelet counts (**C, D**), absolute neutrophil counts (**E, F**), and calprotectin (**G, H**). Spearman’s correlation coefficients were calculated.

### Levels of PAI-1 and tPA associate with severe disease and worse outcomes

Compared with patients breathing room air, patients requiring oxygen had significantly higher levels of PAI-1 (p=0.02) (**Figure 3A**), but not tPA or D-dimer (**Figure 3B-C**). Beyond mode of respiratory support, oxygenation efficiency can also be measured by comparing pulse oximetry (**SpO**_**2**_) to the fraction of inspired oxygen (**FiO**_**2**_). We tested the correlation between PAI-1, D-dimer and SpO_2_/FiO_2_ ratio and found a strong negative association (r=-0.35, p=0.0002 for PAI-1; r=-0.37, p=0.009 for D-dimer;) (**Figure 3D and F**). A negative association was also appreciated between oxygenation efficiency and tPA (r=-0.19, p=0.04), albeit less robust than for PAI-1 and D-dimer (**Figure 3E**). Among the 118 patients, 24 died, 92 were discharged, and two remained hospitalized at the time of this analysis. Significantly higher levels of both PAI-1 (p=0.04) and tPA (p=0.0003) were observed among patients who died as compared with those who were discharged, with this difference being especially robust for tPA (**Figure 3G-H**). Surprisingly, we did not see a significant difference in D-dimer levels between those two groups (**Figure 3I**). In summary, high levels of tPA and PAI-1 were associated with worse respiratory status and poor clinical outcomes; in particular, high levels of tPA were strongly associated with death.

**Figure 3:**
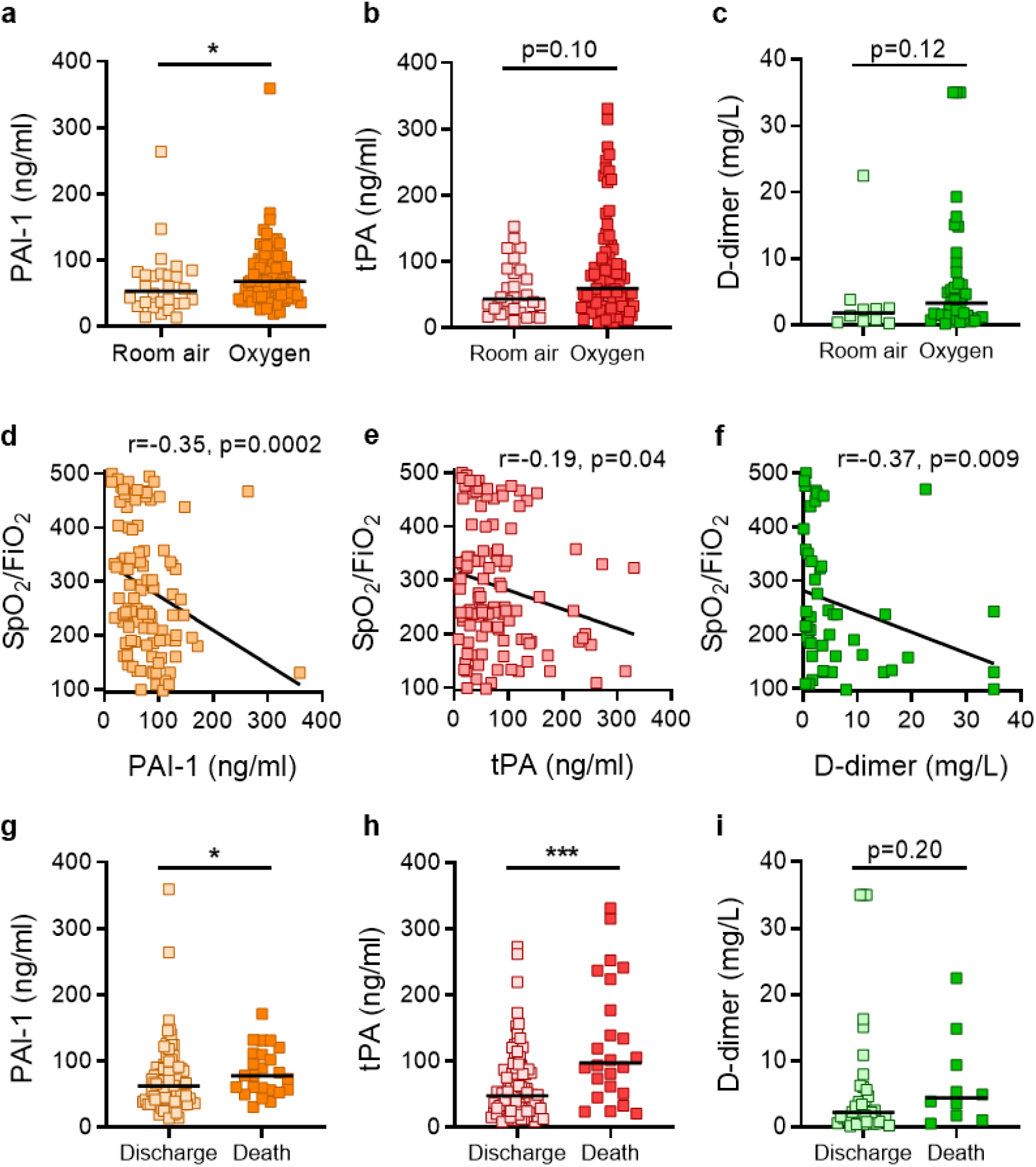
Association between PAI-1, tPA, D-dimer and respiratory status as well as final outcomes. **A-C**, COVID-19 patients were grouped by clinical status (room air vs. supplemental oxygen) and analyzed for PAI-1, tPA, and D-dimer. Level of PAI-1, tPA, and D-dimer were not normally distributed based on Shapiro-Wilk test. Groups were compared by Mann-Whitney test; *p<0.05. **D-F**, PA1-1, tPA, and D-dimer were compared to SpO2/FiO2 ratio for each patient, and correlations were determined by Spearman’s test. **G-I**, COVID-19 patients were also grouped by final outcomes (death vs. discharge). Level of PAI-1, tPA and D-dimer were not normally distributed based on Shapiro-Wilk test. Thus groups were compared by Mann-Whitney test; *p<0.05, ***p<0.001.

### High tPA COVID-19 samples have enhanced spontaneous fibrinolysis

Finally, we asked whether COVID-19 plasma with the highest tPA levels might demonstrate enhanced spontaneous fibrinolysis as compared with low-tPA COVID-19 plasma or control plasma. A spontaneous fibrinolysis assay was performed on 10 COVID-19 plasma samples with high tPA (>100 ng/mL), 10 COVID-19 samples with low tPA (<20 ng/mL), and 10 healthy control plasma samples (mean value 2.4 ng/mL). Notably, the high-tPA COVID-19 samples significantly enhanced spontaneous fibrinolysis as compared with low-tPA and healthy control plasma samples (**Figure 4A-B**). Consistent with this observation, we found that tPA levels were on average 2.2-fold higher than PAI-1 in the high tPA patients (**Supplementary Figure 1A-B**). This was in contrast to the ratio in control plasma samples (where PAI-1 levels averaged more than 10-fold greater than tPA) or in COVID-19 patients with tPA less than 20 ng/mL (where PAI-1 levels were >2-fold greater than tPA). No significant difference in age or oxygenation efficiency were observed in a subset of ten high tPA and ten low tPA patients (**Supplementary Figure 2**). Detailed demographic and clinical characteristics of those COVID-19 patients with high and low tPA are presented in **Supplementary Table 1**.

**Figure 4:**
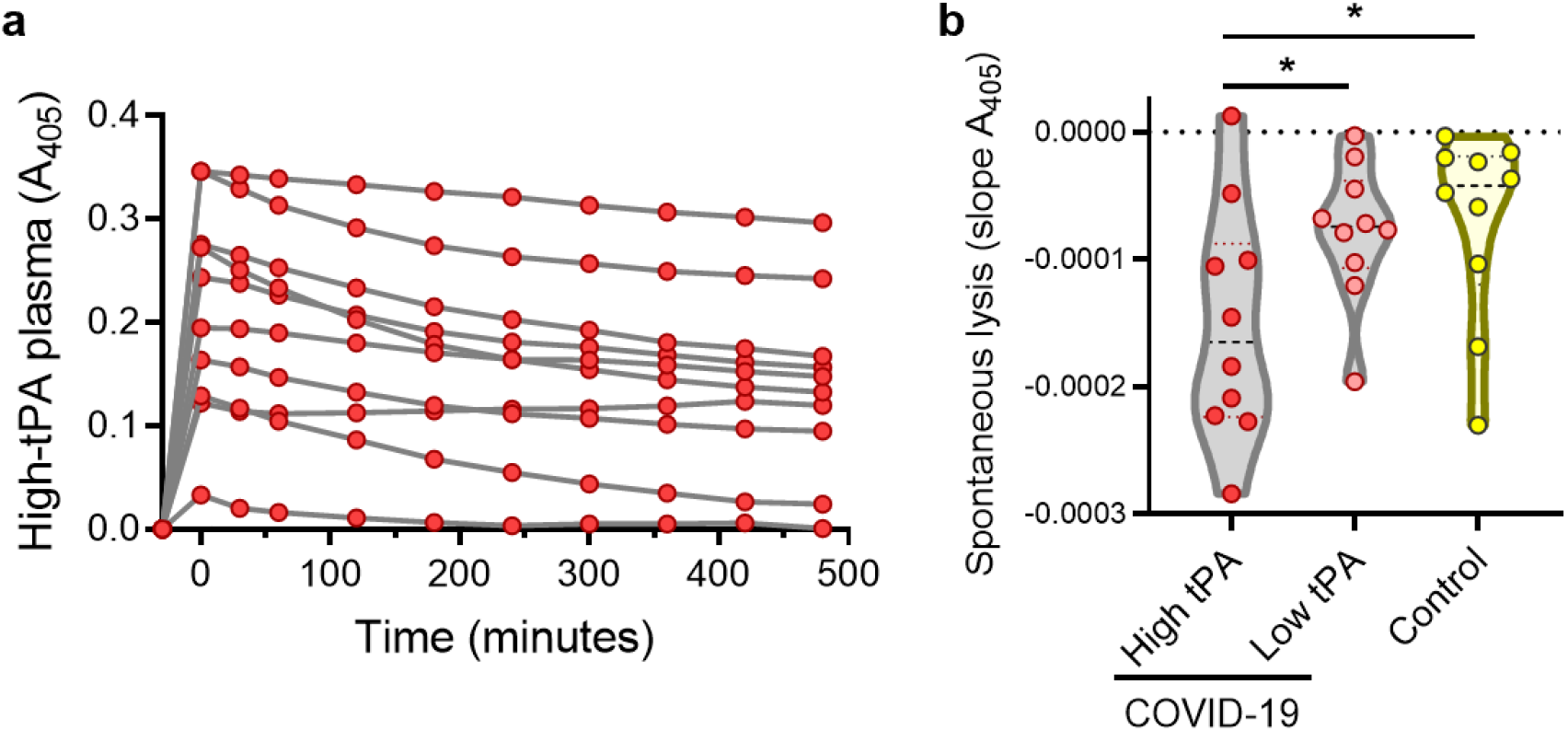
Spontaneous lysis rate among COVID-19 patients with high and low tPA. The ability of COVID-19 patients’ plasma with high (>100ng/mL) and low tPA (<20ng/mL) to promote spontaneous lysis of an *ex vivo* plasma clot formed by the addition of alpha human thrombin was evaluated. **A**. Lysis over time was recorded for 10 high-tPA plasma samples. **B**. The rate of lysis determined from the slope of the absorbance from t_0min_ to t_480min_ was compared between COVID-19 patients with high tPA, low tPA, and healthy controls by one-way ANOVA; *p<0.05.

## DISCUSSION

Fibrinolytic homeostasis in COVID-19 is likely complex and influenced by various factors. Normal lung physiology has a pro-fibrinolytic tendency^25^. However, during acute respiratory distress syndrome (**ARDS**) impaired fibrinolysis results in accumulation of fibrin that promotes hyaline membrane formation and alveolar injury^26^. Fibrin is removed by plasmin. It is believed that depressed fibrinolysis in ARDS is at least partially driven by increased circulating PAI-1 that exerts a negative effect on the plasminogen activation system^25^. Indeed, elevated PAI-1 is an independent risk factor for poor ARDS outcomes^27^. Elevated PAI-1 and its associated hypo-fibrinolytic state were observed in the 2002 SARS-CoV epidemic^28^, while recent characterizations of COVID-19 patients have suggested impaired global fibrinolysis^18,21^. Interestingly, in our large cohort of hospitalized COVID-19 patients, we observed elevated levels of not only PAI-1, but also tPA. While high PAI-1 and D-dimer tracked most closely with impaired oxygenation efficiency, tPA was the best predictor of death. A recent study of 78 hospitalized COVID-19 patients also detected elevations of both PAI-1 and tPA, particularly among critically-ill COVID-19 patients^21^; however, the mechanistic role of the elevated tPA among COVID-19 patients was not specifically investigated. Furthermore, the level of tPA we detected in the COVID-19 patients, 78 ± 68 ng/mL, is striking and much higher than the 23.9 ± 14.5 ng/mL described in this prior report in 48 patients admitted to ICU^21^. Notably these levels are even higher than is observed in trauma patients who have exaggerated fibrinolytic activity and in patients with hantavirus cardiopulmonary syndrome, which carries high alveolar hemorrhage risk^29-31^.

The major source of these high levels of tPA among COVID-19 patients is likely endothelial cells. The source of PAI-1 could also be the endothelium or perhaps release from activated platelets (as we found a strong correlation between PAI-1 and platelet counts). High PAI-1 expression in other cell types such as macrophages has also been reported during hantavirus infections^29^. One hallmark of COVID-19 ARDS is the sequestration of leukocytes, particularly neutrophils, in the microvasculature of the lung—contributing to alveolar injury and unrestricted inflammation^27^. This local proinflammatory environment is further exaggerated by the formation of NETs and results in massive release of proinflammatory cytokines^6^. Those cytokines likely trigger endothelial cell activation and thereby promote local release of tPA and PAI-1^6,32^. Notably, we observed a strong correlation between tPA/PAI-1 and both absolute neutrophil counts and circulating calprotectin, a neutrophil activation marker. In addition to endothelial activation, it is possible that direct infection and destruction of endothelial cells by SARS-CoV-2 may also potentiate the release of tPA and PAI-1^11^.

While the prothrombotic risk associated with COVID-19 is well recognized, the risk of bleeding should not be ignored. One recent large multicenter study observed an overall bleeding risk of 4.8% among hospitalized COVID-19 patients and this risk increased to 7.6% among critically-ill patients^16^. Elevated D-dimer was associated with both thrombotic and bleeding complications^16^. It has been suggested that high PAI-1 levels overcome the effects of local tPA and produce a net prothrombotic hypofibrinolytic state in COVID-19 patients^21^. However, we here found a subset of COVID-19 patients with extremely high levels of tPA (>100ng/mL) in which fibrinolysis seems to dominate. This may at least partially explain the enhanced bleeding risk observed in some groups of patients with COVID-19.

Our study has some limitations. We did not have access to fresh plasma samples each day of a patient’s hospitalization. PAI-1 and tPA levels were therefore not tested on a defined day of hospitalization, but rather when a plasma sample became available to the research laboratory. It should however be noted that when assessing correlations of PAI-1 and tPA with clinical variables, same-day laboratory and clinical status data were used. Due to research restrictions during the pandemic we were not allowed to recruit new healthy controls. Healthy controls were recruited during the pre-COVID-19 era and we were not able to match gender and age to COVID-19 patients. Future studies should endeavor to systematically track PAI-1 and tPA levels over the full course of hospitalization of COVID-19 patients and to compare with gender- and age-matched controls. We also recognize that tPA is not the sole activator of plasminogen, as uPA also plays a role in the fibrinolysis regulation and PAI-1 can also inhibit uPA^17^. Dysregulation of uPA and its receptor system have been implicated in the pathogenesis of pulmonary fibrosis and ARDS^33,34^. The role of uPA and its receptor in COVID-19 warrants further investigation.

Because the COVID-19 associated prothrombotic risk is known, prophylactic anticoagulation has become part of standard COVID-19 treatment. High rates of thromboembolic events from early studies prompted some experts to recommend a more intensive dose of anticoagulation among COVID-19 patients^2^. We would urge caution regarding this recommendation (pending randomized studies) as the coagulopathy of COVID-19 is complex and potentially dynamic. Therapies aimed at promoting fibrinolysis, such as administration of aerosolized or intravenous tPA, have been trialed in ARDS models where there have been some promising preclinical results^35,36^. Profibrinolytic therapy has been suggested as a potential beneficial therapy in COVID-19 patients suffering from ARDS^27^ and is currently being tested in multiple clinical trials (https://clinicaltrials.gov/ct2/results?cond=Covid19&term=tpa). We have now found that a hyperfibrinolytic state exists in some COVID-19 patients. Targeted therapies that promote fibrinolysis therefore need to be selective and cautious to minimize bleeding risk. Finally, our data suggests that high systemic tPA may be a biomarker for poor clinical outcomes and supports further studies of tPA levels during the course of disease progression.

## Data Availability

All data will be made available upon publication.

## ACKNOWLEDGEMENTS

The work was supported by a COVID-19 Cardiovascular Impact Research Ignitor Grant from the Michigan Medicine Frankel Cardiovascular Center (to JSK and YK), the A. Alfred Taubman Medical Research Institute (to JSK and YK), and the National Institutes of Health (HL055374 to DAL). YZ was supported by a career development grant from the Rheumatology Research Foundation. JSK was supported by grants from the NIH (R01HL115138), Lupus Research Alliance, and Burroughs Wellcome Fund. YK was supported by the NHLBI Intramural Research Program, and grants from the NIH (ITAC Award, K08HL131993, R01HL150392),.

## Supplementary Materials

**Supplementary Table 1:** Demographic and clinical characteristics of COVID-19 patients with high and low tPA

|  |  |  |  |  |
| --- | --- | --- | --- | --- |
|  | High tPA |  | Low tPA |  |
| Demographics |  |  |  |  |
| Number | 10 |  | 10 |  |
| Age (years)* | 66 ± 18 | (28-82) | 54 ± 18 | (29-86) |
| Female | 6 | (60%) | 4 | (40%) |
| White/Caucasian | 3 | (30%) | 3 | (30%) |
| Black/African-American | 5 | (50%) | 6 | (60%) |
| Comorbidities |  |  |  |  |
| Diabetes | 3 | (30%) | 1 | (10%) |
| Heart disease | 3 | (30%) | 2 | (20%) |
| Renal disease | 1 | (10%) | 3 | (30%) |
| Lung disease | 0 |  | 4 | (40%) |
| Autoimmune | 1 | (10%) | 2 | (20%) |
| Cancer | 1 | (10%) | 1 | (10%) |
| Obesity | 8 | (80%) | 5 | (50%) |
| Hypertension | 3 | (30%) | 5 | (50%) |
| Immune deficiency | 2 | (20%) | 0 |  |
| History of smoking | 2 | (20%) | 1 | (10%) |
| Medications♦ |  |  |  |  |
| Hydroxychloroquine | 2 | (20%) | 1 | (10%) |
| Anti-IL6 receptor | 1 | (10%) | 1 | (10%) |
| ACE inhibitor | 0 |  | 0 |  |
| Angiotensin receptor blocker | 0 |  | 0 |  |
| Antibiotic | 4 | (40%) | 3 | (30%) |
| Remdesivir | 0 |  | 0 |  |
| Dexamethasone | 0 |  | 0 |  |
| IV heparin | 5 | (50%) | 4 | (40%) |
| Subcutaneous heparin | 2 | (20%) | 3 | (30%) |
| Subcutaneous enoxaparin | 2 | (20%) | 2 | (20%) |
| Alteplase | 0 |  | 0 |  |
| In-hospital thrombosis |  |  |  |  |
| Arterial thrombosis | 0 |  | 0 |  |
| Venous thrombosis | 0 |  | 1 | (10%) |
| Respiratory status♦ |  |  |  |  |
| Room air | 0 |  | 3 | (30%) |
| Nasal Cannula | 2 | (20%) | 4 | (40%) |
| High flow oxygen | 1 | (10%) | 1 | (10%) |
| Mechanical ventilation | 7 | (70%) | 2 | (20%) |
| Awake-prone positioning strategy | 4 | (40%) | 1 | (10%) |
| Final outcomes |  |  |  |  |
| Discharged | 3 | (30%) | 10 | (100%) |
| Death | 6 | (60%) | 0 |  |
| Remains hospitalized | 1 | (10%) | 0 |  |
| * Mean ± standard deviation (range) |  |  |  |  |
| ♦ At time of sample collection |  |  |  |  |

**Supplementary Figure 1.**
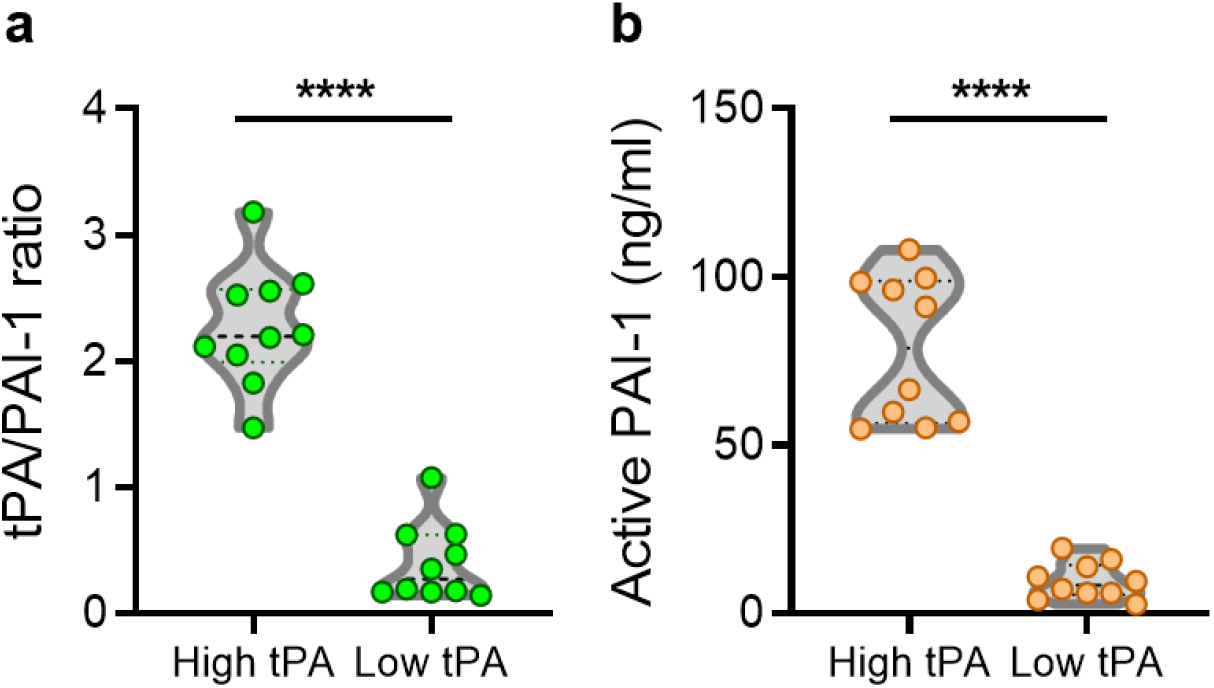
tPA/PAI-1 ratio and PAI-1 activity among COVID-19 patients with high and low tPA. Total tPA and total PAI-1 ratio was determined and active PAI-1 was measured in COVID-19 patients with high tPA (>100ng/mL) or low tPA (<20ng/mL). The ratio of tPA/ PAI-1 (**A**) and levels of active PAI-1 (**B**) were compared by Mann-Whitney test; ****p<0.0001.

**Supplementary Figure 2.**
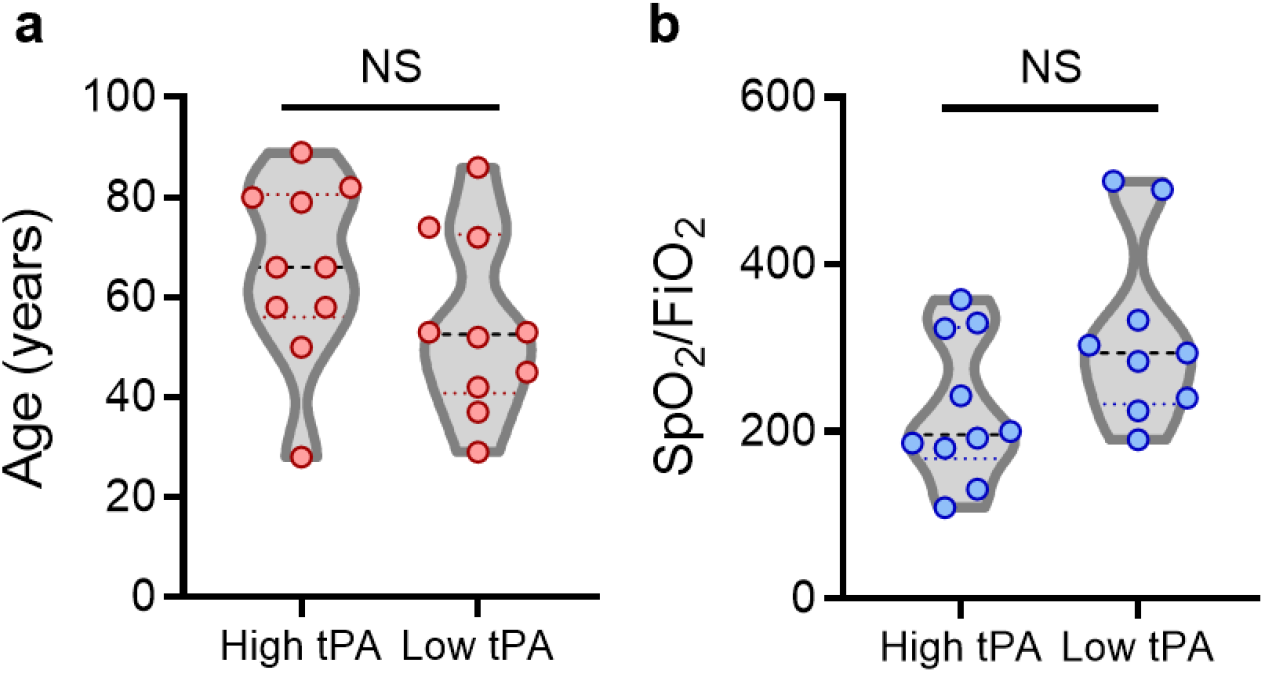
Age and SpO2/FiO2 among COVID-19 patients with high and low tPA. Age (**A**) and SpO2/FiO2 (**B**) were compared by Mann-Whitney test; NS=none significant.

